# Infodemiological study of COVID-19 in Latin America and The Caribbean

**DOI:** 10.1101/2020.08.11.20173054

**Authors:** Carlos Jesús Aragón-Ayala, Julissa Copa-Uscamayta, Luis Herrera, Frank Zela-Coila, Cender Udai Quispe-Juli

## Abstract

**Background:** Infodemiology has been widely used to assess epidemics. In light of the recent pandemic, we use Google Search data to explore online interest about COVID-19 and related topics in 20 countries of Latin America and the Caribbean.

**Method:** Data from Google Trends from 2019/12/30 to 2020/04/25 regarding COVID-19 and other related topics were retrieved and correlated with official data on COVID-19 cases and with national epidemiological indicators.

**Results:** The Latin American and Caribbean countries with the most interest for COVID19 were Peru (100%) and Panama (98.39%). No correlation was found between this interest and national epidemiological indicators. The global and local response time were 20.2±1.2 days and 16.7±15 days, respectively. The duration of public attention was 64.8±12.5 days. The most popular topics related to COVID-19 were: *the country’s situation* (100 ± 0) and *coronavirus symptoms* (36.82±16.16). Most countries showed a strong or moderated (r=0.72) significant correlation between searches related to COVID-19 and daily new cases. In addition, the highest significant lag correlation was found on day 13.35±5.76 (r=0.79).

**Conclusions:** Interest shown by Latin American and Caribbean countries for COVID-19 was high. The degree of online interest in a country does not clearly reflect the magnitude of their epidemiological indicators. The response time and the lag correlation were greater than in European and Asian countries. Little interest was found for preventive measures. Strong correlation between searches for COVID-19 and daily new cases suggests a predictive utility that should be investigated by further studies.

## 1. Introduction

The world is being affected by a disease caused by coronavirus 2019 (COVID-19).

According to the World Health Organization (WHO) the pandemic comes with a rapid and massive diffusion of information, a phenomenon known as infodemic.^1^ Frequently, this information is not correct, causing panic and dangerous behaviors in the population, affecting the effectiveness of public interventions and worsening the situation in low- and middle-income countries. With the aim to fight infodemics, infodemiology rises.

Infodemiology is the science of distribution and determinants of information on the internet or a population, with a final goal to inform about public health and policies.^2,3^ Social network sites and search engines are its main sources of information, Google and Twitter are the most popular.^2^ These web platforms allow a real time analysis,^3^ facilitating outbreak prediction, especially during pandemics.^2^ Further analysis informs about health-related behavior, attitudes and knowledge in the population. ^3^ This serves public health professionals who need to keep close surveillance on quick changes in demand for information whether it is to calm down the population or to detect outbreaks, using the peaks in internet searches as an early predictor.

One of the most reliable tools for infodemiology is Google Search, because of its anonymous and almost universal use. ^3-5^ Google Trends (GT) is a public access tool which analyzes daily searches on Google Search, generating data from a specific time and zone as a Relative Search Volume (RSV).^6^ The use of GT in surveillance of the public interest for an outbreak is promising, because it allows to know the principal queries and doubts around the population. It has been used in studies about infectious diseases, mental health, general diseases, public behavior and its seasonality.^7-11^

Other studies about COVID-19 and GT searches have been conducted in China,^7,12^ Taiwan,^11^ Iran,^13^ The United States of America (USA), United Kingdom (UK), Canada, Ireland, Australia, New Zealand^14^ and Colombia.^15^ With the objective to expand related knowledge in Latin American and Caribbean (LAC) countries, and to improve the previously utilized methods, we analyzed COVID-19 related queries and their relationship with epidemiological indicators_such as daily incidence, lethality, and deaths by COVID-19, for each country belonging to LAC. Additionally, the principal queries related to COVID-19 were determined by country, as well as the response time and duration of the public attention.

## 2. Material and methods

### 2.1. Design and study period

Data from GT^16^ and the Johns Hopkins Center for Systems Science and Engineering (CSSE)^17,18^ was utilized for topics related to COVID-19. The study period was from 30/12/2019 (the day before the first notification to the WHO of disease by COVID-19 in Wuhan) to 25/04/2020, for each one of the 20 LAC countries which were studied (Argentina, Bolivia, Brazil, Chile, Colombia, Costa Rica, Cuba, Ecuador, El Salvador, Guatemala, Honduras, Mexico, Nicaragua, Panama, Paraguay, Peru, Puerto Rico, Dominican Republic, Uruguay and Venezuela). Data was collected from 27/04/2020 to 30/04/2020.

### 2.2. Google Trends

GT allows access to a variety of queries done with the Google engine. Its results are anonymous, categorized (by related searches) and aggregated. This allows it to reflect the worldwide interest on a particular subject.^19,20^. GT normalizes data to ease comparison between terms, as well as normalization on a temporal and geographical level. Normalized data is calculated by dividing the number of searches related to a term by the total of searches done in Google, having previously selected a specific place and time range. Results are scaled from 0 to a 100, resulting in a Relative Search Volume (RSV).^11,14,19-21^ RSV of 100 represents the peak of interest in a particular subject within the chosen area and time, indirectly correcting bias in absolute measurements such as internet access or population size, which varies between countries.^21^ Search terms on GT are not case sensitive, but results can be affected by the use of accentuation. Combined search terms can be created by merging two or more terms with the use of a plus (+). Nonetheless, there is a limited amount of characters which can be used to form these terms. The comparison between terms (simple or combined) has a limit of five.

### 2.3. Search terms

For the general topic of COVID-19 we used the combined term “coronavirus + COVID-19 + SARS-CoV2 + nuevo coronavirus + 2019-nCoV” in Spanish; and: “coronavirus + coronavírus + COVID-19 + SARS-CoV2 + novo coronavirus + novo coronavírus + 2019- nCoV” in Portuguese.

Topics of possible public concern were identified through internet searches, news, newspapers, social network sites, and interviews. Then, for each identified topic, the most popular search terms were found using the tool “Related queries - Top” and “Related queries - Rising”. After several tests, combined search terms were formulated, using terms with the most overall searches. One of the explored topics was a promising treatment, in this case we used Hydroxychloroquine, having the greatest public interest, as well as the largest number of clinical trials^22^ (See **Supplemental file 1**).

### 2.4. Procedures

First, the general term COVID-19 was searched by combining Spanish and Portuguese terms: “coronavirus + COVID-19 + SARS-CoV2 + nuevo coronavirus + 2019-nCoV + novo coronavírus” with “Worldwide” option. Then, a worldwide ranking of average RSV was obtained per country. Data from the LAC countries was extracted, and each average RSV was standardized on a scale from 0 to 100, to facilitate the analysis. Also, for every country, the RSV time series were extracted from the general term of COVID-19 in the study period. Secondly, several comparisons were made for each country, by searching the identified terms in groups of five. This way, searches were ranked in order of popularity using the average RSV for every country. The RSV time series in the study period was extracted for the first five concerns by country.

### 2.5. Global and local response time and duration of public attention

*Global response time* was defined as the time between the day when WHO was notified of the outbreak in Wuhan, and the start of the RSV related to COVID-19.^14^ The *duration of public attention* is the time between the beginning of the RSV of the general terms related to COVID-19 and its maximum peak.^14^. *Local response time* was defined as the time between the date in which the first case in each country and the maximum peak of searches.

### 2.6. Epidemiological data of COVID-19

The CSSE database is publicly available on GitHub.^17,18^. It allows to keep a real-time record of confirmed cases, deaths, and recoveries of COVID-19 from every country, using DXY, a platform directed by the Chinese medical community, as its main data source. Data is updated in a semi-automatic manner, monitoring and requesting information from government websites, as well as the review of social network sites and local news. In addition, it shows proper alignment with the daily reports of the WHO and the Chinese Center for Disease Control and Prevention.^18^

New cases and daily deaths were calculated with the accumulated data. In this way, the epidemiological indicators analyzed in this study were: total confirmed cases, average new daily cases, average incidence (per 100 habitants), total deaths, mortality (per 100 000 habitants), lethality rate ([cumulative deaths / cumulative confirmed cases] * 100) and the percentage of total confirmed cases by total population. United Nations total population data by country were used.^23^

### 2.7. Statistical analysis

The GT data was downloaded in a CSV (comma-separated values) format which was then imported into Microsoft Excel 2019. The RSV values reported as “<1”, were replaced by 0.5 to facilitate analysis. Analysis and graphs were done in STATA 14. A linear time series analysis of the RSV related to COVID-19 is presented, as well as new cases, daily deaths, and main concerns. Furthermore, scatterplot graphs were made utilizing the average RSV by country and its epidemiological indicator.

Spearman’s correlation coefficient was calculated (because of the non-normality of the data), considering weak (r ≤ 0.35), moderate (0.36 - 0.67) and strong (0.68 - 1.0)^24^ and considering p <0.05, significant. It was calculated between the average RSV related to COVID-19 per country and its epidemiological indicator. Spearman’s analysis of lag correlation (p <0.05) was performed between the number of daily new cases and the daily RSV on COVID-19 per country, to determine if an increase in the RSV is correlated with a subsequent increase in new cases of COVID-19, as previous studies.^11,25^

### 2.8. Ethical considerations

This study was conducted guided by the principles of scientific and ethical integrity. Its approval by an ethics committee was not necessary because the data used is anonymous and freely accessible.

## 3. Results

### 3.1. Interest in COVID-19 in Latin American and Caribbean countries

The highest public interest (expressed as average RSV) in COVID-19 was shown by Peru (100%), followed by Panama (98.39%), Colombia (83.65%), Paraguay (82.65%), Uruguay (80.65%), Argentina (80.65%), Bolivia (77.42%), Ecuador (75.81%), Costa Rica (72.58%), Chile (70.97%), Guatemala (70.97%), Venezuela (66.13%), Mexico (66.13%), Dominican Republic (58.06%), Brazil (46.77%), Cuba (1.61%), El Salvador (1.61%), Honduras (1.61%), Nicaragua (1.61%) and Puerto Rico (1.61%). The 90% of South American countries and 50% of Central American countries (Mexico, Dominican Republic, Guatemala, Costa Rica, and Panama) have an average RSV over 50%.

### 3.2. Relationship between epidemiological indicators and interest about COVID-19

No statistically significant correlation was found between the average RSV of each country and epidemiological indicators such as: Percentage of the total population that presented COVID-19, average incidence per 100 000 habitants, observed lethality rate, total confirmed cases, average daily new cases, accumulated deaths, and deaths per 100,000 habitants. See **Figure 1**.

**Figure 1.**
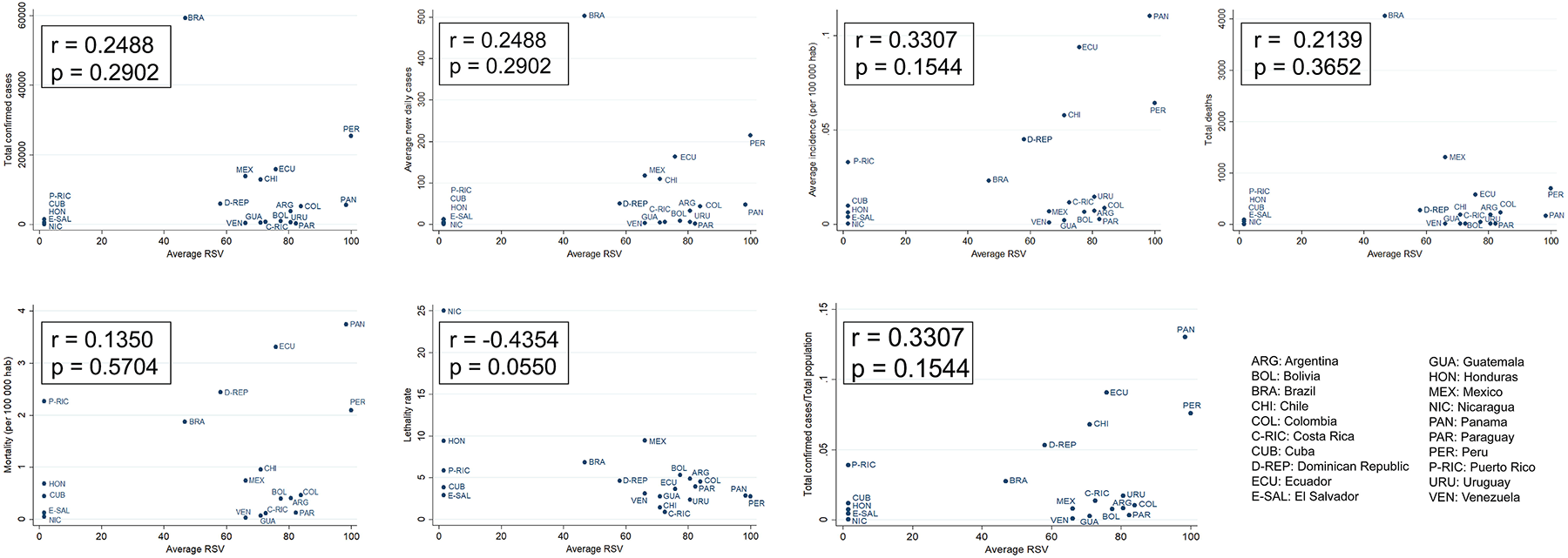
Relative Search Volume for COVID-19 and epidemiological indicators in Latin American and Caribbean countries. Data from 2019/12/30 to 2020/04/25. r: Spearman’s rank correlation coefficient, p: p-value.

### 3.3. Trends of interest for COVID-19 and its relationship with daily new cases and deaths

In most countries, the presence of three main search peaks with variable magnitude were observed approximately on January 30 (± 18 days), February 26 (± 8 days) and March 19 (± 10 days), respectively. The average magnitude of these peaks is around 18%, 25% and 100% for the first, second, and third peak respectively. Furthermore, a general decrease in searches can be observed in most countries. For more details, see **Figure 2**.

**Figure 2.**
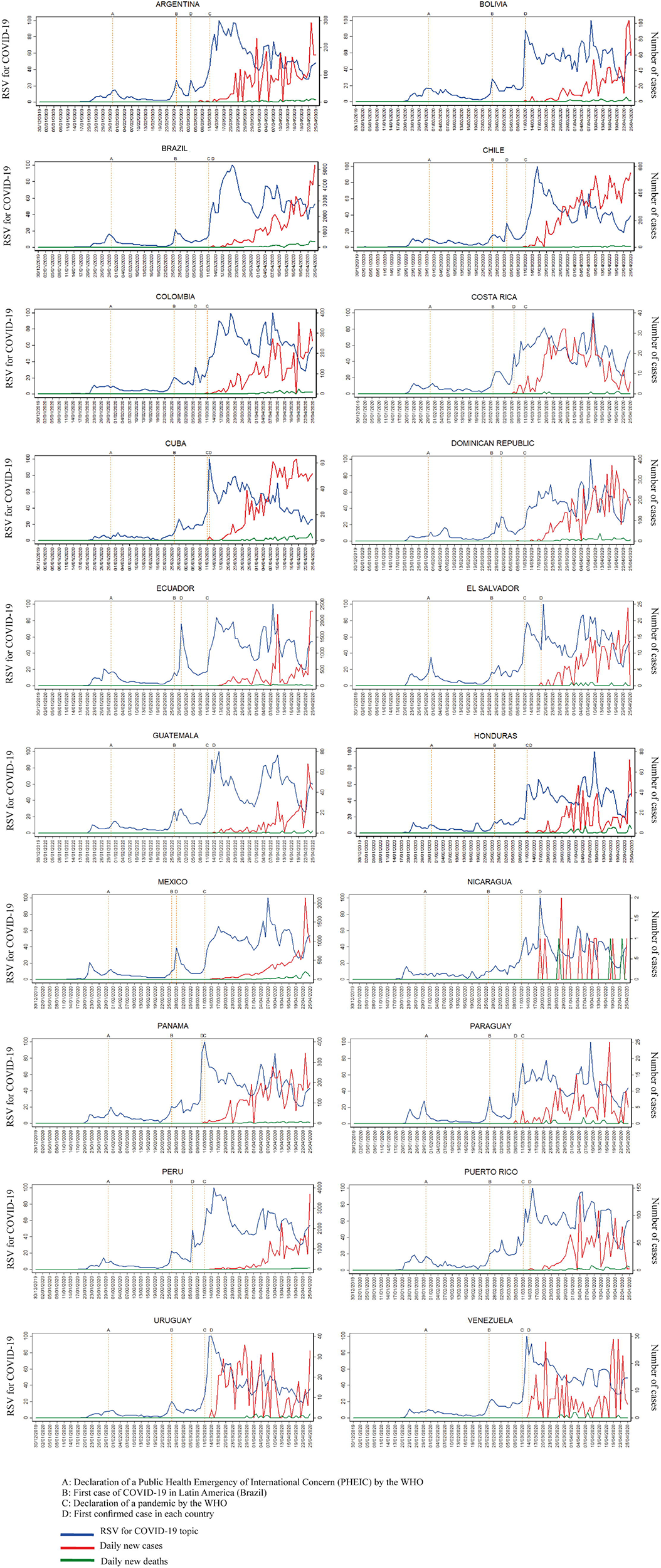
Relative Search Volume for COVID-19, daily number of new cases and daily number of deaths by COVID-19 in Latin American and Caribbean countries. Data from 2019/12/30 to 2020/04/25.

### 3.4. Global and local response time, and duration of public attention

On average, the global and local response time were 20.2 ± 1.2 days and 16.7 ± 15 days, respectively. The duration of public attention was 64.8 ± 12.5 days. The local response time was negative in Uruguay and Venezuela. See **Figure 3** and **Supplemental file 2**

**Figure 3.**
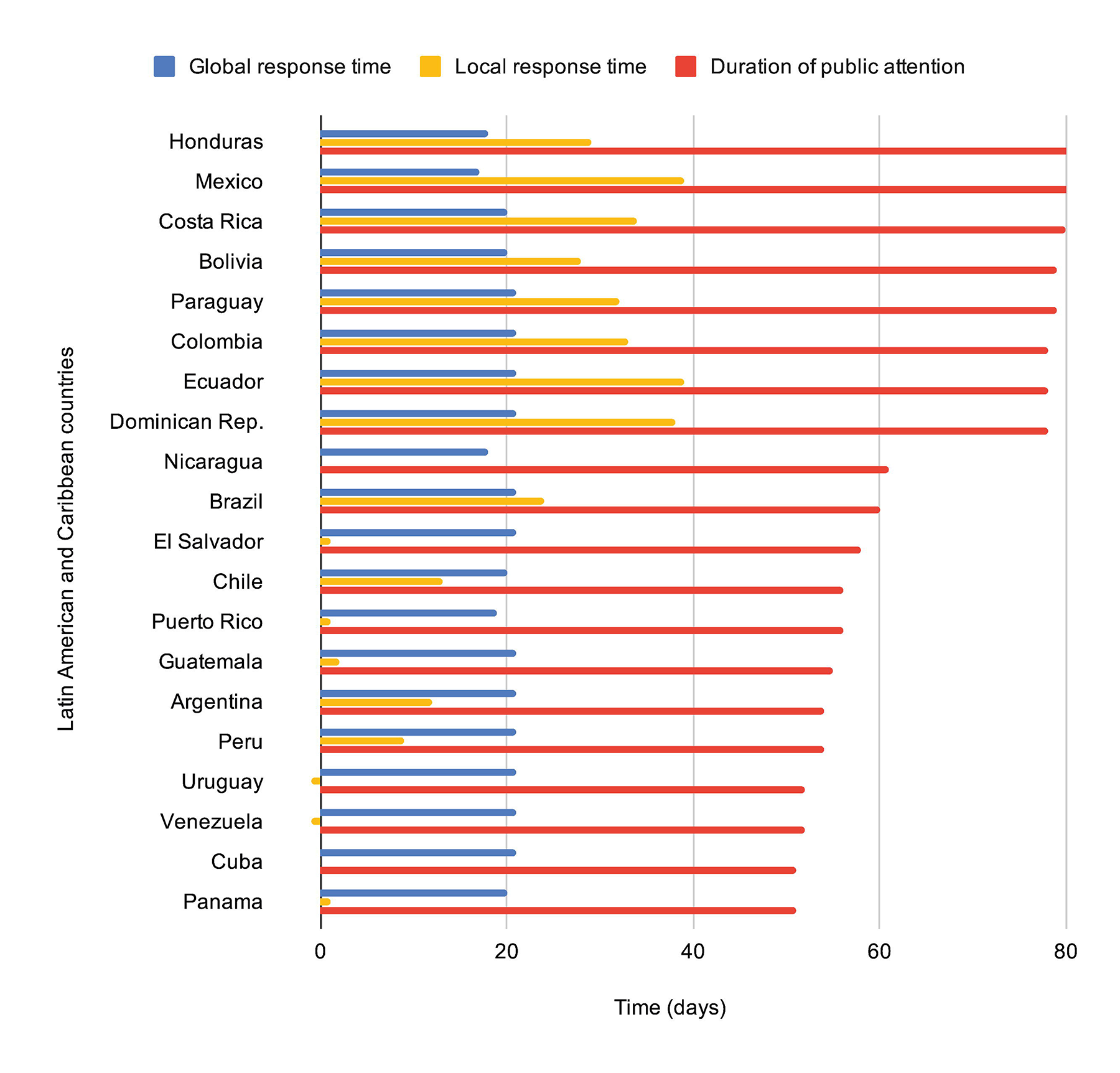
Global and local response time and duration of public attention regarding to COVID-19 in Latin American and Caribbean countries. Data from 2019/12/30 to 2020/04/25.

### 3.5. Interest in topics related to COVID-19 per country

The main interested topics related to COVID-19 were *the country’s situation regarding coronaviruses* (100.00 ± 0.00), *coronavirus symptoms* (36.82 ± 16.16), *mask* (23.55 ± 16.43), *coronavirus definition* (18.71 ± 10.60), *quarantine and social isolation* (10.69 ± 6.92). Additionally, the other subjects were of less interest on average: *vaccine* (5.72 ± 2.35), *hand sanitizer* (5.60 ± 6.85), *coronavirus prevention* (5.32 ± 3.85), *hand washing* (3.46 ± 4.38), *medical treatment* (3.40 ± 1.79), *coronavirus transmission* (3.04 ± 1.53), *coronavirus tests* (1.90 ± 1.23), *coronavirus origin* (1.79 ± 1.14), *attention to coronavirus* (1.64 ± 0.82), *risk population* (0.86 ± 0.89), *cleaning and disinfection against coronavirus* (0.51 ± 0.31), promising treatment (Hydroxychloroquine) (0.41 ± 0.35), *home or natural remedies* (0.28 ± 0.31), *mental health* (0.23 ± 0.19). **Table 1** presents these concerns for each country. The top 10 of the most used terms in the study period can be seen in **Supplemental file 3**.

**Table 1.** Average Relative Search Volume for topics related to COVID-19 in Latin American and Caribbean countries

| Topics related to COVID-19 | Latin American and Caribbean countries |  |  |  |  |  |  |  |  |  |  |  |  |  |  |  |  |  |  |  |
| --- | --- | --- | --- | --- | --- | --- | --- | --- | --- | --- | --- | --- | --- | --- | --- | --- | --- | --- | --- | --- |
|  | Argentina | Bolivia | Brazil | Chile | Colombia | Costa Rica | Cuba | Ecuador | El Salvador | Guatemala | Honduras | Mexico | Nicaragua | Panama | Paraguay | Peru | Puerto Rico | Dominican Republic | Uruguay | Venezuela |
| Coronavirus definition | 21.82 | 21.21 | 13.38 | 7.27 | 9.52 | 9.28 | 1.18 | 13.69 | 20.00 | 16.29 | 27.74 | 20.84 | 31.25 | 12.70 | 25.00 | 11.24 | 22.39 | 39.27 | 8.39 | 41.67 |
| Symptoms | 30.00 | 33.33 | 43.48 | 27.78 | 19.05 | 37.50 | 5.88 | 36.84 | 46.67 | 23.53 | 44.44 | 41.67 | 55.56 | 35.71 | 40.00 | 21.43 | 54.84 | 81.82 | 23.08 | 33.85 |
| Country's situation about COVID-19 | 100.00 | 100.00 | 100.00 | 100.00 | 100.00 | 100.00 | 100.00 | 100.00 | 100.00 | 100.00 | 100.00 | 100.00 | 100.00 | 100.00 | 100.00 | 100.00 | 100.00 | 100.00 | 100.00 | 100.00 |
| Coronavirus transmission | 6.35 | 4.71 | 1.11 | 3.43 | 1.50 | 2.11 | 0.41 | 2.51 | 2.67 | 1.73 | 3.03 | 2.58 | 5.56 | 4.10 | 4.75 | 2.38 | 2.84 | 3.43 | 1.60 | 4.09 |
| Coronavirus prevention | 6.01 | 6.29 | 7.52 | 1.84 | 1.50 | 2.41 | 0.73 | 3.14 | 4.00 | 4.16 | 11.09 | 3.85 | 11.11 | 4.10 | 6.21 | 4.08 | 3.16 | 6.51 | 2.20 | 16.41 |
| Mask | 32.73 | 21.21 | 66.67 | 1.73 | 6.67 | 14.58 | 1.31 | 23.95 | 33.33 | 23.53 | 29.63 | 41.67 | 33.33 | 21.43 | 9.00 | 12.71 | 38.39 | 40.91 | 6.87 | 11.28 |
| Hand washing | 3.34 | 3.65 | 0.73 | 21.16 | 2.11 | 3.22 | 0.10 | 2.51 | 3.33 | 1.49 | 1.75 | 3.85 | 4.44 | 2.46 | 5.46 | 3.50 | 2.63 | 1.54 | 0.62 | 1.28 |
| Hand sanitizer | 15.87 | 7.07 | 26.76 | 1.51 | 0.33 | 9.28 | 0.00 | 0.96 | 13.33 | 4.99 | 1.89 | 1.29 | 7.11 | 1.23 | 10.00 | 1.88 | 0.24 | 0.75 | 6.87 | 0.56 |
| Vaccine | 7.93 | 7.07 | 7.52 | 4.00 | 3.86 | 3.62 | 2.94 | 4.89 | 6.67 | 3.46 | 8.07 | 3.85 | 11.11 | 6.93 | 5.14 | 3.18 | 4.68 | 10.20 | 4.90 | 4.41 |
| Quarantine, and social isolation | 24.55 | 24.24 | 2.63 | 9.26 | 11.43 | 4.02 | 1.10 | 7.33 | 20.00 | 5.82 | 8.07 | 8.33 | 3.33 | 14.29 | 5.00 | 13.87 | 10.66 | 14.73 | 6.87 | 18.23 |
| Medical treatment | 2.66 | 4.38 | 3.76 | 1.84 | 1.35 | 1.25 | 2.61 | 2.20 | 4.00 | 2.89 | 6.05 | 2.37 | 6.67 | 3.46 | 2.92 | 1.34 | 5.74 | 7.06 | 1.60 | 3.83 |
| Home or natural remedies | 0.15 | 0.33 | 0.01 | 0.12 | 0.14 | 0.01 | 0.02 | 0.35 | 0.24 | 0.09 | 0.23 | 0.07 | 0.18 | 0.20 | 0.58 | 0.11 | 1.22 | 0.88 | 0.14 | 0.49 |
| Cleaning and disinfection against coronavirus | 1.12 | 1.29 | 0.31 | 0.49 | 0.33 | 0.35 | 0.00 | 0.55 | 0.53 | 0.15 | 0.23 | 0.33 | 0.89 | 0.41 | 0.70 | 0.38 | 0.55 | 0.63 | 0.62 | 0.42 |
| Attention to coronavirus | 2.97 | 1.45 | 1.34 | 1.36 | 1.58 | 1.41 | 0.13 | 1.13 | 1.78 | 1.63 | 0.87 | 1.29 | 1.78 | 1.54 | 1.26 | 1.14 | 3.51 | 3.08 | 2.64 | 0.90 |
| Mental health | 0.24 | 0.17 | 0.25 | 0.19 | 0.10 | 0.05 | 0.05 | 0.26 | 0.36 | 0.15 | 0.12 | 0.14 | 0.35 | 0.20 | 0.28 | 0.13 | 0.97 | 0.16 | 0.18 | 0.23 |
| High-risk population | 4.01 | 0.92 | 2.23 | 0.99 | 0.31 | 0.69 | 0.11 | 0.62 | 0.24 | 0.20 | 0.35 | 0.20 | 1.11 | 0.35 | 0.63 | 0.94 | 0.73 | 0.63 | 1.25 | 0.70 |
| Coronavirus origin | 2.38 | 3.65 | 0.92 | 0.72 | 0.69 | 1.13 | 0.20 | 1.22 | 2.29 | 1.63 | 3.26 | 1.08 | 3.33 | 1.76 | 1.64 | 0.83 | 1.62 | 2.31 | 0.78 | 4.41 |
| Coronavirus tests | 0.56 | 1.60 | 3.01 | 2.64 | 0.94 | 2.11 | 0.25 | 1.57 | 1.42 | 1.40 | 1.89 | 2.08 | 2.22 | 2.31 | 0.63 | 1.73 | 5.30 | 4.34 | 0.93 | 1.10 |
| Promising treatment (Hydroxychloroquine) | 0.40 | 1.10 | 0.73 | 0.26 | 0.11 | 0.05 | 0.01 | 0.55 | 0.21 | 0.05 | 0.52 | 0.12 | 1.11 | 0.13 | 0.47 | 0.44 | 0.36 | 1.10 | 0.35 | 0.14 |

### 3.6. Trends of the 5 main topics related to COVID-19 by countries

In most countries there was a first peak of searches near the 30^th^ January for topics about *the country’s situation, symptoms, definition*, and *prevention*, while searches for *face masks, quarantine, hand-washing* and *gel alcohol* remain constant. However, since the last week of February, there was a first irregular increase in most searches (highest peak for *the country ‘s situation* and *symptoms*) and a later decrease in other searches, with some exceptions. (See **Figure 4**)

**Figure 4.**
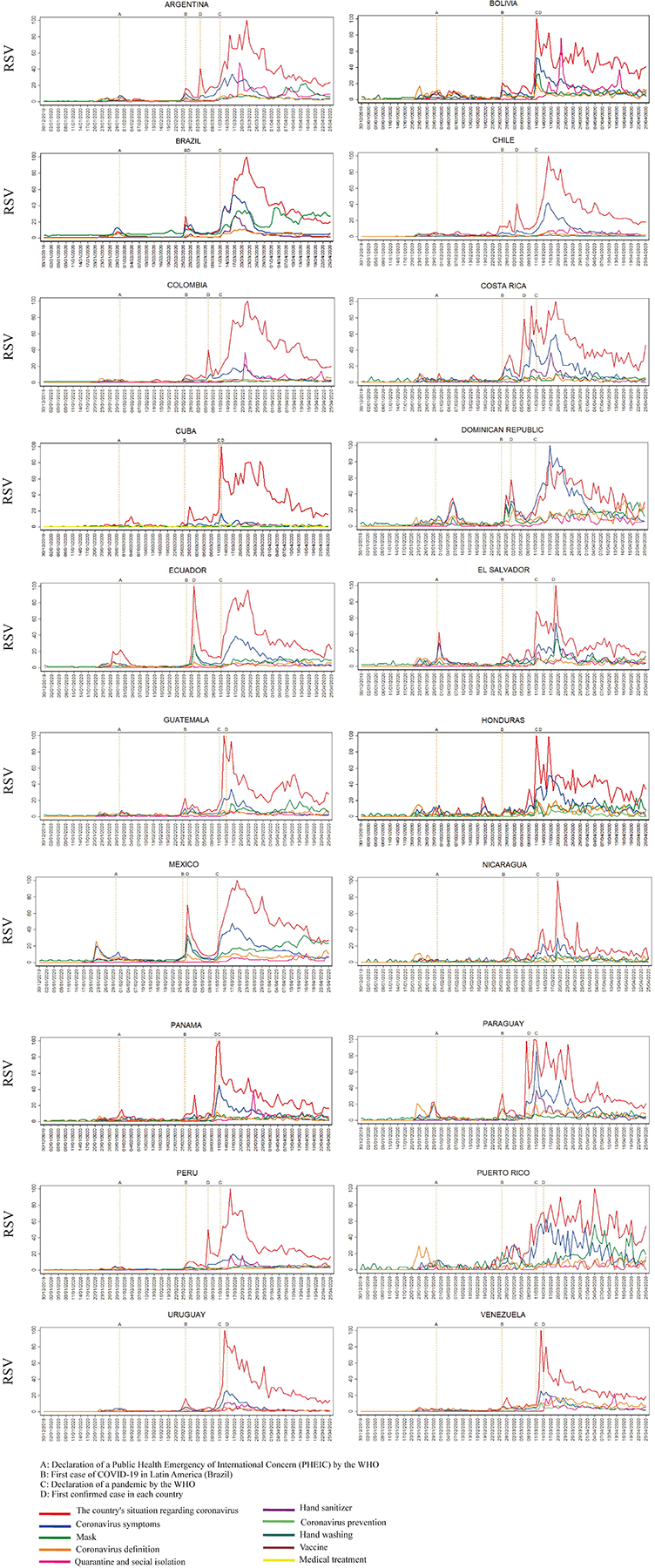
Relative Search Volume for top five topics related to COVID-19 in Latin American and Caribbean countries. Data from 2019/12/30 to 2020/04/25.

### 3.7. Lag correlation between daily new cases and internet queries for topic regarding COVID-19

The correlation between daily new cases and RSV for COVID-19 topic was significant (p < 0.001) and strong in most countries (r ~ 0.72), moderate in El Salvador (r = 0.61) and weak in Nicaragua (r = 0.35). In the lag period, all correlations were significant (p <0.001 in all countries, except for Nicaragua with a maximum p = 0.0161), with a high correlation (r = 0.79) 5.76 ± 13.35 days earlier for daily new cases (See **Figure 5** and **Table 2**).

**Figure 5.**
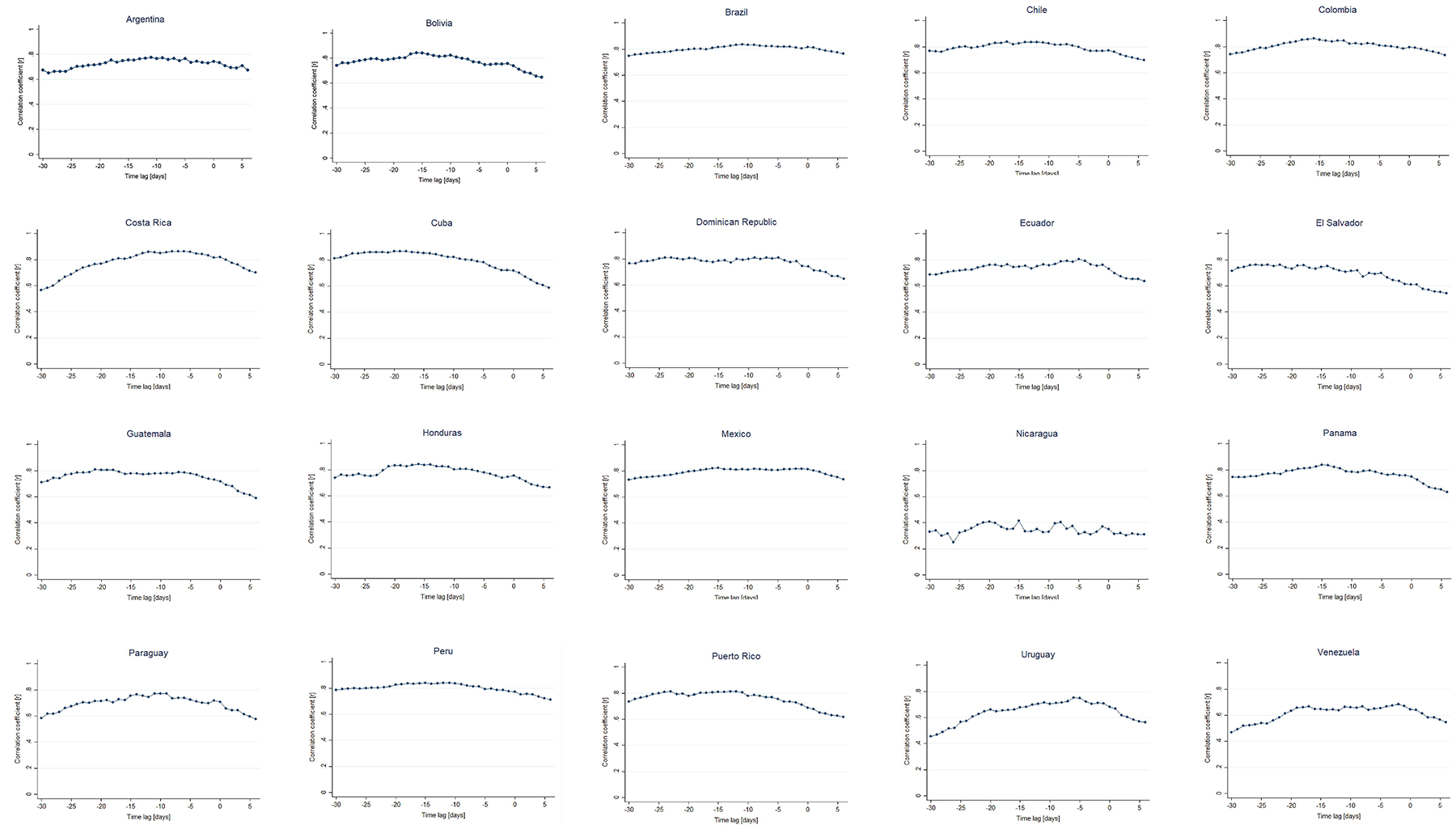
Lag correlations of Relative Search Volume for COVID-19 and daily new cases in Latin American and Caribbean countries. Data from 2019/12/30 to 2020/04/25.

**Table 2.**
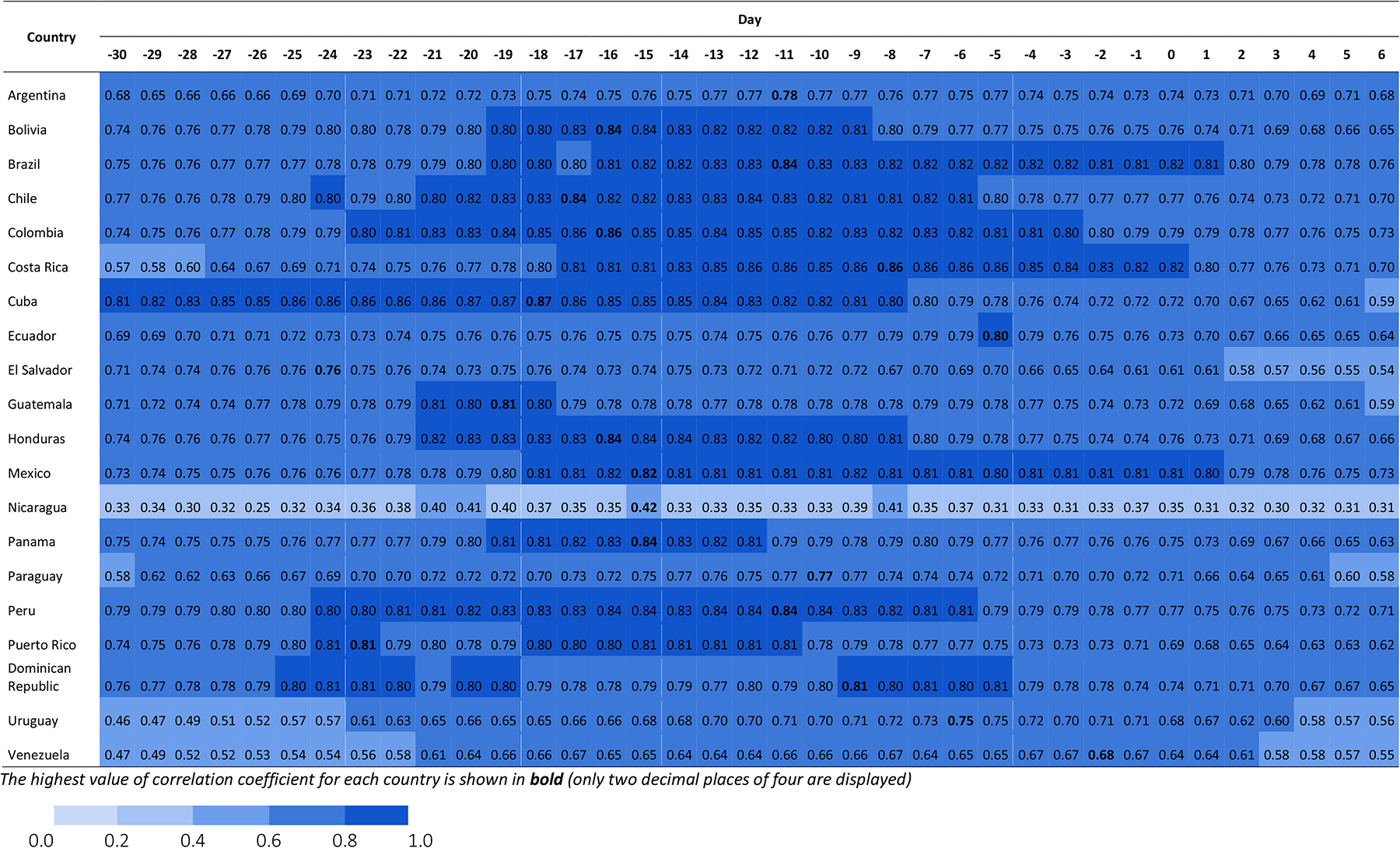
Lag correlations coefficient of Relative Search Volume for COVID-19 and daily new cases in Latin American and Caribbean countries

## 4. Discussion

### 4.1. Interest about COVID-19 in Latin American and Caribbean countries, and its relationship with epidemiological indicators

The majority of LAC countries showed a high interest for COVID-19, except Cuba, El Salvador, Honduras, Nicaragua and Puerto Rico. They showed an interest of approximately 2% of Peru (the country with the highest interest). In a previous study, interest about COVID- 19 was higher in countries with the most confirmed cases at that moment (Italy, France and Spain).^12^ This suggests that the number of confirmed cases would be a factor related to popular interest for COVID-19. However, we found no significant correlation between epidemiological indicators (such as total confirmed cases) and the average relative interest by country. It is possible that intervening factors or external causes of this relationship were not explored, thus not allowed to find it. Some factors could be the proportion of the population with internet access, dissemination of these indicators through official and unofficial media, popularity and use of different sources of information, spread of news via social network sites, concern for national and international reality, Human Development Index, and others.^14,26^ Future studies should investigate the role of these and other factors to better understand the applicability of different tools such as Google Trends, in LAC countries.

### 4.2. Trends of interest in COVID-19 and its relationship with daily new cases and deaths

The three major peaks in searches observed, in most countries, could be explained by their proximity to high impact national and international events. The first peak would be related to the declaration of a Public Health Emergency of International Concern (PHEIC) by the WHO (January 30, 2020)^27^ and the declaration of national alerts by health authorities in countries such as Colombia.^15^ This is consistent with findings in USA, UK, Canada, Ireland, New Zealand, Australia,^14^ China,^7,12,28^ Taiwan,^11^ Republic of Korea, Japan, Germany, Iran, Italy,^29.31^ and Spain,^28^ which also showed an increase in their searches near that date. The second main peak occurred around the confirmation of the first case of COVID-19 in Latin America that took place in Brazil (February 26, 2020).^32^ This could be explained by the impact of the notification of cases of a country in the interest and the searches of neighboring countries, as in nearby states of the USA.^33^ Future studies should deeply analyze the impact of notable events in each country on their epidemiological indicators for a better comprehension. The third peak would correspond to the declaration of a pandemic by the WHO (March 11, 2020),^27^ the confirmation of the first cases and deaths, and the establishment of mitigation or suppression policies during March and April in each country, which is consistent with previous studies.^11,14,28,31,33,35^ A trend of progressive decrease in interest in COVID-19 is also observed in most countries, even despite the increase in cases and deaths, similar to prior investigations.^12,28,29,33,35^ This limits the use of Google Trends as an epidemiological surveillance tool in the long term, as has been suggested in background studies.^7,13,15,29,31,36^ This can be explained by an increase in the global awareness due to an effective national dissemination,^12^ or a decrease in the interest of people over time, similar to viral news of rapid rise and rapid fall.^14,28,29,37^ However, particular features of each country would impact the trends evolution, therefore, further research is needed.

### 4.3. Global and local response time and duration of public attention

LAC countries began their searches for COVID-19 approximately 20 days after the declared outbreak in Wuhan, similar to Ireland and New Zealand which started 21 days later,^14^ but different from the USA, UK, Canada and Australia which their searches began 3, 10, 14, and 10 days later respectively.^14^ This difference might be explained by the confirmation of cases in neighboring countries, because in our context, the first case in the Americas region was confirmed on January 21 (USA). The duration of public attention was highly variable between countries, but on average (65 days) was higher than the USA, UK, Canada, Ireland, New Zealand and Australia, which presented 27, 23, 13, 12, 11 and 22 days, respectively.^14^ It is possible that a shorter attention time favors the spread of the outbreak.^14^ However, there would be an intrinsic dynamism according to the period analyzed. The local response time was on average 17 days but it was close to zero in Cuba, El Salvador, Guatemala, Nicaragua, Panama, Puerto Rico, Uruguay and Venezuela. This could be due to the impact of the first confirmed cases on the public interest, even contributing to the low number of confirmed cases in these countries, although it will be necessary to test this hypothesis in forthcoming studies.

### 4.4. Interest in topics related to COVID-19 and Trends of the top 5 topics related to COVID-19

The main topics related to COVID-19 with the highest average interest in Latin America countries were: *the country’s situation regarding COVID-19, symptoms, mask, definition of coronavirus, quarantine*, and *social isolation*. This is comparable to the USA, UK, Canada, Australia and New Zealand, where searches about situation in the country, definition of coronavirus and its symptoms were predominant,^14,33^ unlike Iran^13^ or Taiwan^11^ where topics related to prevention, such as hand washing and use of antiseptics, were the most popular. This way, despite the differences between countries, there are common concerns, highlighting symptoms, but showing a lesser interest for preventive measures, at least in the early stages.^33^ The evolution of trends follows a variable pattern by country. Some countries such as Bolivia and Mexico had a rapid growth in all their searches, including preventive measures. In others, preventive measures occurred days or weeks later, and to a lesser extent. In Brazil, searches about masks remain relatively constant and in moderate magnitude, different from other countries that showed lower interest. This could be due to effective communication interventions by the health authorities, as a consequence the population did not need to search for additional information on the internet, explaining the decrease in interest. Another explanation could be ineffective interventions, thus generating a constant search trend.

### 4.5. Lag correlation between interest on COVID-19 and confirmed cases

A strong and positive correlation was found between searches for COVID-19 and the subsequent increase in the number of daily confirmed new cases after approximately 14 days. However, the correlation was moderate and weak in El Salvador and Nicaragua, respectively. One possible explanation is that the regions with the lowest correlation have less access to the internet.^11^ Compared with other studies, this lag time and the magnitude of correlation was greater or similar than other countries such as Taiwan (r = 0.70) from 1 to 3 days,^11^ China (r = 0.96) from 5 to 10 days,^7^ Germany, Austria, Australia, Japan, Egypt, UK (r ~ 0.70) with 11.5 days on average.^29^ Furthermore, the instantaneous correlation (lag-0) in most LAC countries was strong (r ~ 0.75), similar to other studies in Australia, Austria, Germany, Egypt, Japan, UK (r ~ 0.60), Spain, Italy,^31^ but different from countries such as the USA, UK, Canada, Ireland, Australia, and New Zealand, which presented weak and moderate correlations (r ~ 0.4).^14^ This variable correlation across studies could be explained by the fact that some studies used an accumulated number of cases for lag correlation analysis, but others used a number of daily new cases.^29^ Another investigation, in Italy, found a correlation using more specific terms such as “cough” or “fever” symptoms, compared to general symptoms.^38^ Although this suggests that interest of people is a possible predictor of first cases of an outbreak in each country,^7^ future studies should control other factors to elucidate in a better way the predictive value of GT searches. Our findings give an idea that the LAC population had more time to obtain information about COVID-19, a period of time that should have been used by governments because in these circumstances people are more sensitive, participative and receptive to information.^13^

### 4.6. Limitations

Our study, like other studies using GT, has some limitations. We used only one data source (GT), it could generate a possible selection bias. The study units were the countries; therefore, important subnational factors could not be evaluated. The representativeness of the sample studied cannot be guaranteed due to the heterogeneous internet access in LAC countries. The complexity and dynamism of epidemiological data diminish external validity at the future stages of COVID-19 pandemic. There is a possibility that the selection of the search terms is incomplete due to linguistic complexity and variability, and cultural worldview of each country, despite having carried out a systematic process of identification of terms. In addition, our results cannot be replicated because the algorithm of GT is not publicly accessible and it could change over time.^33^

### 4.7. Strengths

Despite our limitations, this is the first study that analyzes and explores the quantitative and qualitative relationship of epidemiological indicators of COVID-19 and the concerns of population on this subject, with the largest number of countries studied until now (20 countries) and from the same geographical region (Latin America and the Caribbean). In addition, we used several terms related to topics regarding COVID-19, having previously verified that they were the most used through multiple tests, unlike studies in China (7), Taiwan,^11^ among others, that used one or a few terms, and even other studies did not mention or make the terms used accessible,^14,15^ making it difficult to analyze the validity and replicability of their results, as mentioned in methodological GT studies.^4,5^ Furthermore, to our knowledge, this is the first study to compare the average interest in COVID-19 and its relationship with several epidemiological indicators in each country, simulating an ecological analysis.

### 4.8. Recommendations

Future studies should explore the correlation between adjusted epidemiological data and other search terms. Additionally, other data sources such as social network sites, national call centers for COVID-19, and the media should be added to the analysis.

## 5. Conclusions

Google Trends can be used to know the current status and evolution of the queries, thoughts, concerns, and needs of the study population in multiple periods. Health-policy makers could utilize these brief moments of active searching of the population to provide relevant information for either preventive reasons or to fight infodemics.^1^ Our findings indicate the need that health authorities provide information in a massive way days before the confirmation of cases in the country, because people would be looking for information at that time. In this way, authorities can anticipate infodemics, which in many cases distort reality, inducing panic and dangerous practices, which are deeply rooted in public awareness despite future valid information. The correlation found between the number of new cases and interest for COVID-19 could set a framework for future predictive models. The high response time values in LAC countries compared to high-income countries, gives us an idea of the low awareness about an international issue, which would go unnoticed unless it became a very close threat.

In summary, interest for COVID-19 in Latin America and the Caribbean is high in most countries. Its national trends were influenced by international events, such as the first case reported in the USA and Brazil, or the declaration of the outbreak as a Public Health Emergency of International Importance, then declared as a pandemic by the WHO, as well as national events, for instance, first confirmed cases and deaths in its territory or in neighboring countries, and the implementation of mitigation or suppression policies to a lesser extent. However, the degree of interest for COVID-19 does not clearly reflect the magnitude of epidemiological indicators in a country. The response time and the lag correlation in Latin America and the Caribbean were greater than European and Asian countries, suggesting that they had more time to provide information, failing to take an advantage of this opportunity. It is remarkable that searches in most LAC countries have focused little on preventive measures. In most countries, there is a strong correlation between searches for COVID-19 and the number of daily new cases, therefore, the number of searches could serve as a predictive indicator of new cases, although future studies are necessary to prove this hypothesis. This is the first infodemiological study to approach most of the countries in a continent, this is why we recommend continuing with more studies with a macroregional focus.

## Data Availability

Our raw data is publicly available in Figshare repository: https://doi.org/10.6084/m9.figshare.12644222

## Supplemental files

Supplemental file 1. Search terms for topics related to COVID-19 in Spanish and Portuguese

Supplemental file 2. Global and local response time and duration of public attention regarding COVID-19 in Latin American and Caribbean countries

Supplemental file 3. Top 10 ranking relative queries regarding COVID-19 in Latin American and Caribbean countries

